# Social Determinants of Health and Healthcare Utilization Disparities among Older Adults with and Without Cognitive Impairment

**DOI:** 10.1101/2024.07.14.24310385

**Authors:** Zahra Rahemi, Sophia Z. Shalhout, Juanita-Dawne R. Bacsu, Darina V. Petrovsky, Preeti Pushpalata Zanwar, Swann Arp Adams

## Abstract

The purpose of this study was to determine the healthcare utilization patterns in a national sample of older adults across several social determinants of health factors (ethnicity, gender, race, education) with normal and dementia/impaired cognition. We used datasets from the Health and Retirement Study (HRS, 2018) to evaluate healthcare utilization, including metrics such as hospital and nursing home stays, hospice care, and number of visits to the doctor. Logistic models were used to predict healthcare utilization separately in those with normal cognition and dementia. Our final sample comprised 15,607 adults (mean age: 65.2 normal cognition, mean age 71.5 dementia). Hispanics with normal cognition were less likely to stay in a hospital than non-Hispanic respondents (OR: 0.52-0.71, p<0.01). Being female was associated with a higher risk for shorter nursing home days (OR: 1.41, p<0.01) and doctor visits (OR: 1.63-2, p<0.01) in cognitively normal older adults. Being female was associated with a lower risk for hospital stay in those with dementia (OR: 0.50-0.78, p<0.01). Respondents identifying as Black or other races with dementia were less likely to experience nursing home days (OR: 0.42, p<0.04). Black respondents with normal cognition were less likely to experience doctor visits (OR: 0.32-0.37, p<0.01). Those with more than a high school education in both groups were more likely to experience doctors’ visits. The study points to the continued disparities in healthcare utilization linked to participants’ social determinants of health factors and cognition.

---

Cognitive impairment due to dementia has become a pressing public health concern. An estimated 6.7 million Americans aged 65 and older live with Alzheimer’s disease and related dementia (ADRD) (Alzheimer’s Association, 2024). This number is projected to more than double by 2060 (Rajan et al., 2021), disproportionately affecting minority groups, including Black, Hispanic, and underserved older adults. For example, Hispanic individuals are about 1.5 times more likely to develop ADRD than non-Hispanic Whites (Alzheimer’s Association, 2024). Research indicates that Black individuals aged 50 years and older are two to three times more likely than Caucasians to develop dementia (Garcia et al., 2019). In addition, racial and ethnic minorities typically experience suboptimal quality in care, limited access to healthcare (Mcmaughan et al., 2020), and structural inequities (Peterson et al., 2024) while requiring more frequent and complex healthcare services. This may potentially result in higher rates of healthcare utilization and increased medical costs (Charron-Chénier & Mueller, 2018; Pereira et al., 2022). These findings highlight the crucial need for research to tackle the growing challenges in healthcare utilization and disparities among diverse older adults with cognitive impairment (Pereira et al., 2022; Tappen et al., 2017).

Multiple factors, including social determinants of health (SDOH) such as gender, race, ethnicity, age, education, employment, health literacy, insurance coverage, and resource availability, along with the intersections between these factors, often contribute to disparities in access to and utilization of healthcare services (Chen et al., 2016; Rahemi, 2019; Rahemi & Williams, 2020). Additionally, rural communities often experience unique barriers to accessing cognitive healthcare compounded by limited public transportation, healthcare specialists, finances, and education, as well as challenges related to geographic distance (Bacsu et al., 2019). Greenwood-Ericksen and Kocher note that rural areas have a higher rate of unnecessary emergency department visits, partly due to a lack of healthcare professionals. Other factors like waiting too long to seek care, cultural beliefs, and feeling ashamed can also make it harder for people, especially women, to receive the right care (Greenwood-Ericksen & Kocher, 2019; Wiese et al., 2023). As a result, disadvantaged individuals often experience poor health outcomes, exacerbating systemic healthcare disparities (Charron-Chénier & Mueller, 2018; Wiese et al., 2023).

Healthcare costs are notably higher for racial minority individuals with ADRD compared to minority individuals with normal cognition and White individuals with and without ADRD (Cooper et al., 2010; Pereira et al., 2022). Additionally, older adults with ADRD exhibit higher rates of emergency department visits, hospitalizations, and prolonged hospital stays than their counterparts without ADRD (Benner et al., 2018; Hunt et al., 2018; Kent et al., 2019). The increased rates of healthcare utilization, in turn are associated with increased mortality rates, higher risks of fall-related injuries, earlier admissions to nursing homes, and changes in both cognitive and physical health (Colligan et al., 2017; Pereira et al., 2022). To address these disparities, some scholars emphasize the role of advance care planning (ACP), which involves proactively communicating healthcare decisions before a person loses decision-making capacity (McMahan et al., 2021). Utilizing the past 2014 HRS dataset, we previously demonstrated that participants who were younger, Hispanic, Black, had lower levels of education, or resided in rural areas were less likely to complete ACP. (BLINDED FOR REVIEW). Furthermore, those individuals with ACP were more likely to experience longer stays in hospitals, nursing homes, and home healthcare (Rahemi, Malatyali, et al., 2023).

Research is essential to tackle critical SDOH, including race, ethnicity, sex, and gender, and the role of advance care planning as well as their intersections. These factors contribute to the increasing healthcare challenges and disparities observed in diverse older adults, particularly those with cognitive impairment. Research in this field concerning ADRD and cognitive disorders is limited (Hinton et al., 2024; Pereira et al., 2022). A recent review of 96 randomized controlled trials to support cognition related to dementia (n= 37,278) identified that only 39 trials (39.4%) included ethnicity and only 11.4% (95% CI, 7.5 to 15.9%) of all participants were non Caucasian (Vyas et al., 2018). However, as the American population becomes more racially and ethnically diverse, there is a critical need for research to examine disparities in healthcare utilization for diverse people living with ADRD and cognitive disorders (Hinton et al., 2024).

To gain a more comprehensive understanding of the disparities in care for ADRD, we analyzed healthcare utilization patterns in a national sample of older adults. Our focus was on key SDOH, including age, gender, race, ethnicity, education, marital status, and rural location. We further sought to determine whether any SDOH associations found with the analysis were mediated by the ACP measures of having a living will, durable power of attorney for healthcare, or both. We utilized Health and Retirement Study (HRS, 2018) datasets to evaluate healthcare utilization among a diverse population of U.S. older adults. This involved analyzing data on the total length of hospital stays, nursing home stays, hospice care, and the frequency of doctor visits over the past two years.

## Materials and Methods

Our observational, cross-sectional study utilized data from the Health and Retirement Survey (HRS), a nationally representative survey of respondents aged 51 years and older (Sonnega et al., 2014). The survey utilizes a probability sampling method to ensure a representative demographic sample, with an oversampling of African American, Hispanic, and Floridian participants. For our study, we analyzed a sample of 15,607 respondents from the HRS 2018 survey (wave 14). Our analysis incorporated data from the harmonized HRS version B pertaining to end-of-life topics and the 2018 Rand HRS Longitudinal version 2. As our research utilized secondary data analysis of de-identified data, it was exempt from review by the university’s Institutional Review Board.

### Measures

For analysis, respondents were categorized into two groups using the Langa-Weir approach: dementia/impaired cognition (scores of 1-11) and normal cognition (scores of 12 or higher) (Langa et al., 2017). Because the distributions for all the healthcare utilization variables were highly skewed, we ultimately grouped these variables into three or four categories: never, low, moderate, and high utilization (hospital stay and doctor visits) or never, some, and high utilization (nursing home and hospice care).

Our literature review could find no commonly used cut points; therefore, we based our cut points on natural breaks in the frequency distribution of each variable. We aimed to align these points across variables to the greatest extent possible. Table 1 details the division and cut points for each healthcare variable. Using a data-driven approach, we inspected histograms and box plots to identify these breaks and employed gap statistics to determine the optimal number of bins or clusters, ensuring significant gaps between clusters.

**Table 1.** Healthcare utilization categories and their cut points.

| Utilization Type | Never | Low<br>(# of days) | Medium<br>(# of days) | High<br>(# of days) |
| --- | --- | --- | --- | --- |
| Hospital Days | 0 | 1-7 | 8-15 | 16+ |
| Doctor Visits | 0 | 1-6 (visits) | 8-25 (visits) | 26+ (visits) |
|  | Never | Some | High |  |
| Nursing Home | 0 | 1-119 | 120+ |  |
| Hospice Care | 0 | 1-15 | 16+ |  |

For the duration of hospitalization within the preceding two years, the groups were never (0), low (1-7 days), moderate (8-15 days), and high (16+ days). For nursing home care in the previous two years, the cut points were never (0), low (1-30 days), moderate (31-199 days), and high (120+ days). The number of doctor visits in the previous two years collapsed into never (0), low (1-6 visits), moderate (8-25 visits), and high (26+ visits).

Rurality was based upon the patient’s residence-rural or urban. For the ACP mediation analysis, living will and durable power of attorney for healthcare were captured as present (yes/no). For the combination variable, respondents were classified as no ACP measure (neither living will or durable power of attorney for healthcare), at least one ACP measure (either living will or durable power of attorney for healthcare), or having both ACP measures.

### Statistical Analysis

All analyses were performed using SAS v9.4. An alpha level of less than or equal to 0.05 was set for statistical significance. Given the potential for bias from the differing healthcare needs of individuals with dementia or impaired cognition compared to those with normal cognition. All analyses were stratified by cognition group as described previously. Due to the complex sampling design of the HRS, participant sample weights were applied to all descriptive statistics to ensure nationally representative estimates. Weighted frequencies and means were calculated for the appropriate variables. Weighted sample t-tests and Rao-Scott chi-square tests were employed to determine significant differences between cognition groups (dementia/impaired cognition versus normal cognition).

Previous analyses have shown that using sample weights in multivariable modeling can introduce bias into the estimates (Winship & Radbill, 1994); therefore, we did not incorporate these weights in any of the model results presented. The Proc logistic function was used with the glogit option to conduct polytomous logistic regression models. Each healthcare utilization variable was included as a dependent variable and demographic variables were incorporated as independent variables. The ‘never’ utilization was the reference group for each model. All polytomous logistic models were stratified by cognition group (dementia/impaired cognition versus normal cognition) to facilitate the comparison of various predictors.

For the mediation analysis, we employed the Baron and Kenny method (Baron & Kenny, 1986). Initially, we assessed whether each ACP variable was associated with the SDOH variable and the healthcare utilization outcome. If both conditions were met, a single model was constructed, including the advance care planning variable, the relevant SDOH variable, and the healthcare outcome variable as the dependent variable. Mediation was considered to exist if the previously significant SDOH variable no longer achieved statistical significance in this combined model.

## Results

Table 2 describes and compares the demographic characteristics of the dementia/impaired cognition group in relation to the normal cognition group. In contrast to the normal cognition group, the dementia/impaired cognition group participants had significantly less education, were more likely to be from a racial/ethnic minority community, single, and live in a rural area.

**Table 2.** Descriptive Statistics for the 2018 Heath Retirement Survey Cohort* By Cognition Group (HRS, 2018).

|  | Normal Cognition<br>N=12,540<br>Weighted % (n) | Dementia/Impaired Cognition<br>N=3067<br>Weighted % (n) | P-Value |
| --- | --- | --- | --- |
| Education |  |  |  |
| ≤ 12 years | 36.1 (5215) | 69.4 (2254) | < 0.01 |
| 13+ years | 63.9 (7374) | 30.6 (825) |  |
| Sex |  |  |  |
| Male | 45.9 (6503) | 45.9 (1297) | 0.93 |
| Female | 54.1 (9165) | 54.1 (1782) |  |
| Race |  |  |  |
| White | 82.3 (8784) | 63.6 (1591) | < 0.01 |
| Black/Other | 17.7 (3756) | 36.4 (1476) |  |
| Ethnicity |  |  |  |
| Hispanic | 9.0 (1811) | 18.2 (699) | < 0.01 |
| Non-Hispanic | 91.0 (10759) | 81.8 (2371) |  |
| Marital Status |  |  |  |
| Married/Living with a Partner | 67.0 (7829) | 49.0 (1496) | <0.01 |
| Single/Widowed | 33.0(4760) | 51.0 (1583) |  |
| Rurality |  |  |  |
| Rural | 31.4 (3672) | 36.2 (1034) | <0.01 |
| Urban | 68.6 (8917) | 63.8 (2045) |  |
| Hospitalization Days Categories |  |  |  |
| None (0 days) | 79.9 (9762) | 71.4 (2151) | <0.01 |
| Low (1-7 days) | 15.7 (2081) | 20.0 (579) |  |
| Medium (7-15 days) | 2.6 (383) | 4.7 (143) |  |
| High (16+ days) | 1.8 (252) | 3.9 (110) |  |
| Hospice Care Days Categories† |  |  |  |
| None (0 days) | 79.2 (3724) | 80.5 (4217) | 0.05 |
| Some (1-15 days) | 13.4 (629) | 13.3 (694) |  |
| High (16+ days) | 7.4 (347) | 6.2 (323) |  |
| Nursing Home Days Categories |  |  |  |
| None (0 days) | 97.9 (12237) | 94.7 (2858) | <0.01 |
| Some (1-119 days) | 1.7 (222) | 3.2 (103) |  |
| High (120+ days) | 0.4 (49) | 2.1 (53) |  |
| Doctors Visits Categories |  |  |  |
| None (0 visits) | 8.9 (1168) | 16.9 (522) | < 0.01 |
| Low (1-6 visits) | 51.9 (6022) | 48.3 (1252) |  |
| Medium (7-25 visits) | 33.7 (4076) | 28.9 (742) |  |
| High (26+ visits) | 5.5 (644) | 5.9 (143) |  |

|  | **Mean (SE)** | **Mean (SE)** |  |
| --- | --- | --- | --- |
| Age (yrs) | 65.2 (0.2) | 71.5 (0.4) | < 0.01 |

\* Sampling weights are not provided for deceased participants so only alive respondents are included for weighted analyses in this table.
† This variable is derived from the exit interview survey of deceased participants, which does not have sample weights. Thus, variance estimates have not been adjusted for the sampling design.

Compared to the normal cognition group, those in the dementia/impaired cognition group utilized a greater proportion of hospital and nursing home care and were more likely to have fewer low and medium doctor visits and increased high and no doctor visits. In the dementia/impaired cognition group, the mean age was significantly greater than the normal cognition group (71.5 compared to 65.2).

### Hospital Stay

In the normal cognition group, when examining each level of hospitalization to those who had never been hospitalized, Hispanics were significantly less likely to have low, moderate, and high hospital stays compared to non-Hispanic participants. Older age was significantly associated with hospitalization at each level of utilization.

Among those with dementia/impaired cognition, when examining each level of hospital use, Hispanics were significantly less likely to utilize low and moderate lengths of stay compared to the non-Hispanic group. Females were significantly less likely to have hospital stays. Older age was associated with a significant trend going from short to long stays; however, only the point estimate for the short hospital stays reached statistical significance, with older age demonstrating a greater likelihood of a short hospital stay (Table 3).

**Table 3.**
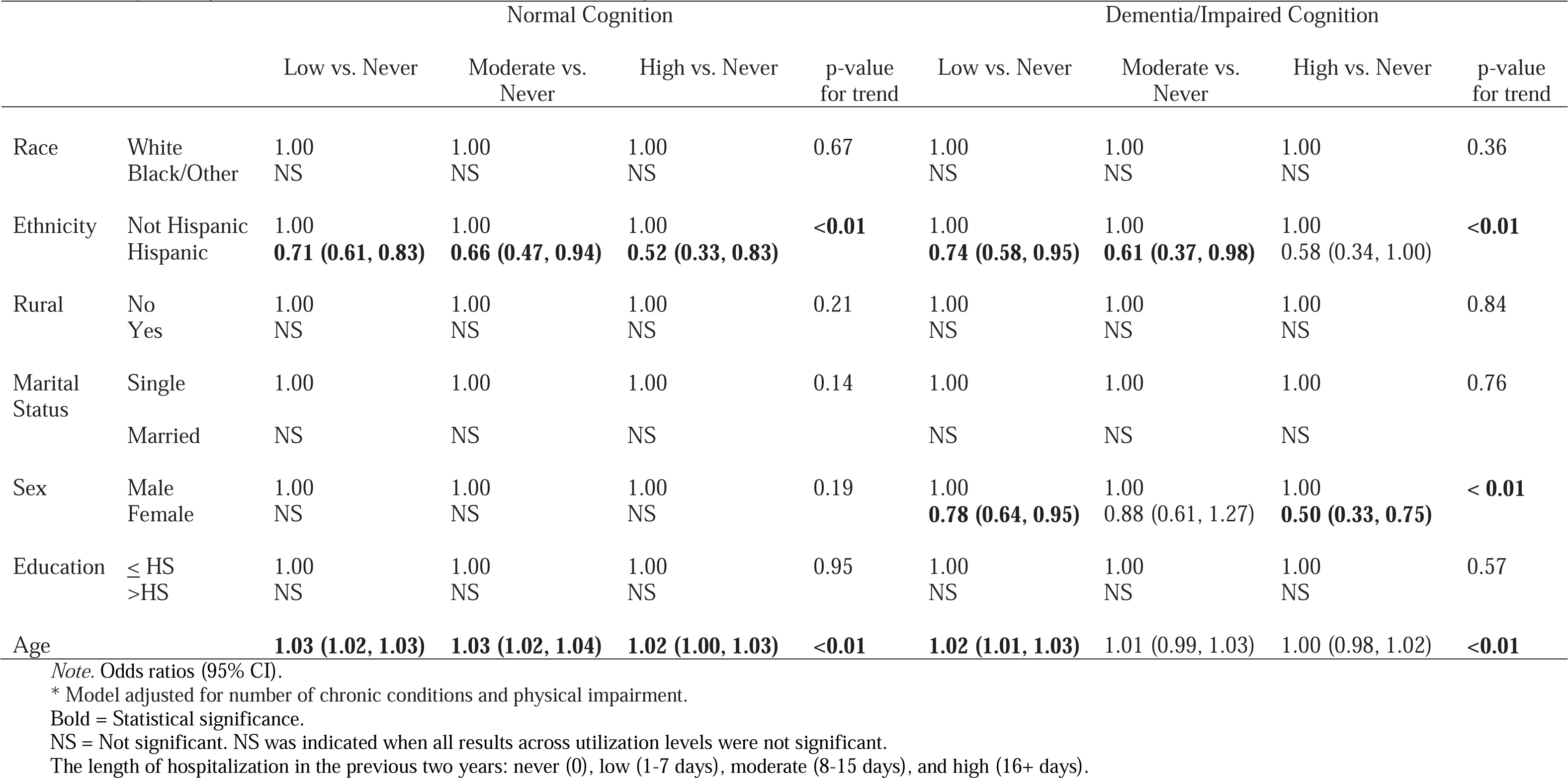
Hospital Days Prediction Model*, Health Retirement Survey, 2018.

|  |  | Normal Cognition |  |  |  | Dementia/Impaired Cognition |  |  |  |
| --- | --- | --- | --- | --- | --- | --- | --- | --- | --- |
|  |  | Low vs. Never | Moderate vs. Never | High vs. Never | p-value for trend | Low vs. Never | Moderate vs. Never | High vs. Never | p-value for trend |
| Race | White | 1.00 | 1.00 | 1.00 | 0.67 | 1.00 | 1.00 | 1.00 | 0.36 |
|  | Black/Other | NS | NS | NS |  | NS | NS | NS |  |
| Ethnicity | Not Hispanic | 1.00 | 1.00 | 1.00 | <0.01 | 1.00 | 1.00 | 1.00 | <0.01 |
|  | Hispanic | **0.71 (0.61, 0.83)** | **0.66 (0.47, 0.94)** | **0.52 (0.33, 0.83)** |  | **0.74 (0.58, 0.95)** | **0.61 (0.37, 0.98)** | 0.58 (0.34, 1.00) |  |
| Rural | No | 1.00 | 1.00 | 1.00 | 0.21 | 1.00 | 1.00 | 1.00 | 0.84 |
|  | Yes | NS | NS | NS |  | NS | NS | NS |  |
| Marital Status | Single | 1.00 | 1.00 | 1.00 | 0.14 | 1.00 | 1.00 | 1.00 | 0.76 |
|  | Married | NS | NS | NS |  | NS | NS | NS |  |
| Sex | Male | 1.00 | 1.00 | 1.00 | 0.19 | 1.00 | 1.00 | 1.00 | < 0.01 |
|  | Female | NS | NS | NS |  | **0.78 (0.64, 0.95)** | 0.88 (0.61, 1.27) | **0.50 (0.33, 0.75)** |  |
| Education | ≤ HS | 1.00 | 1.00 | 1.00 | 0.95 | 1.00 | 1.00 | 1.00 | 0.57 |
|  | >HS | NS | NS | NS |  | NS | NS | NS |  |
| Age |  | **1.03 (1.02, 1.03)** | **1.03 (1.02, 1.04)** | **1.02 (1.00, 1.03)** | **<0.01** | **1.02 (1.01, 1.03)** | 1.01 (0.99, 1.03) | 1.00 (0.98, 1.02) | **<0.01** |

Note. Odds ratios (95% CI).
\* Model adjusted for number of chronic conditions and physical impairment.
Bold = Statistical significance.
NS = Not significant. NS was indicated when all results across utilization levels were not significant.
The length of hospitalization in the previous two years: never (0), low (1-7 days), moderate (8-15 days), and high (16+ days).

### Nursing Home Stay

In the normal cognition group, when examining each level of stay, married individuals were significantly less likely to utilize all lengths of nursing home stays, while older participants were more likely to utilize all levels of nursing home stays. Women were significantly more likely to experience shorter stays in the nursing home while significantly less likely to utilize longer stays (see Table 3- trend test). Hispanic participants were significantly less likely to utilize some nursing home care compared to non-Hispanic individuals (Table 4).

**Table 4.** Nursing Home Days Prediction Model*, Health Retirement Survey, 2018.

|  |  | Normal Cognition |  |  | Dementia/Impaired Cognition |  |  |
| --- | --- | --- | --- | --- | --- | --- | --- |
|  |  | Some vs. Never | High vs. Never | p-value<br>for trend | Some vs. Never | High vs. Never | p-value<br>for trend |
| Race | White | 1.00 | 1.00 | 0.06 | 1.00 | 1.00 | <**0.04** |
|  | Black/Other | NS | NS |  | 0.75 (0.48, 1.18) | **0.42 (0.21, 0.88)** |  |
| Ethnicity | Not Hispanic | 1.00 | 1.00 | **0.03** | 1.00 | 1.00 | <**0.01** |
|  | Hispanic | **0.46 (0.26, 0.83)** | 1.00 (0.34, 2.96) |  | **0.23 (0.10, 0.52)** | **0.25 (0.07, 0.82)** |  |
| Rural | No | 1.00 | 1.00 | 0.52 | 1.00 | 1.00 | 0.20 |
|  | Yes | NS | NS |  | NS | NS |  |
| Marital Status | Single | 1.00 | 1.00 | <**0.01** | 1.00 | 1.00 | <**0.01** |
|  | Married | **0.64 (0.47, 0.85)** | **0.23 (0.11, 0.48)** |  | 0.85 (0.55, 1.31) | **0.26 (0.12, 0.57)** |  |
| Sex | Male | 1.00 | 1.00 | <**0.01** | 1.00 | 1.00 | 0.20 |
|  | Female | **1.41 (1.03, 1.92)** | **0.49 (0.26, 0.91)** |  | NS | NS |  |
| Education | ≤ HS | 1.00 | 1.00 | 0.11 | 1.00 | 1.00 | 0.81 |
|  | >HS | NS | NS |  | NS | NS |  |
| Age |  | **1.05 (1.03, 1.06)** | **1.10 (1.07, 1.13)** | <**0.01** | **1.04 (1.02, 1.06)** | **1.07 (1.04, 1.10)** | <**0.01** |

Note. Odds ratios (95% CI).
\* Model adjusted for the number of chronic conditions and physical impairment.
Bold = Statistical significance.
NS = Not significant. NS was indicated when all results across utilization levels were not significant.
Nursing home care in the previous two years: never (0), some (1-199 days), and high (120+ days).

Among dementia/impaired cognition and normal cognition groups, marital status (p<0.01) and age had a similar trend effect on nursing home utilization: lower utilization for married and higher utilization for older individuals. In contrast to the normal cognition group, in the dementia/impaired cognition group, race and ethnicity also proved to have a significant trend in utilization, while rurality and sex did not have a significant trend. Based upon the point estimate testing, those of Black/other race and Hispanic ethnicity in dementia/impaired cognition group were significantly less likely to utilize high lengths of nursing home stays. Hispanic participants were significantly less likely to utilize some nursing home care compared to non-Hispanic individuals.

### Hospice Care

Among the normal cognition group, only increased age was associated with a significantly increased likelihood of utilizing hospice care (Table 5). Among those with dementia/impaired cognition, individuals who had more education were significantly more likely to have moderate utilization compared to less educated individuals. Furthermore, with increasing age, individuals with impaired cognition or dementia were more likely to utilize a longer length of hospice care (see Table 4- trend test).

**Table 5.**
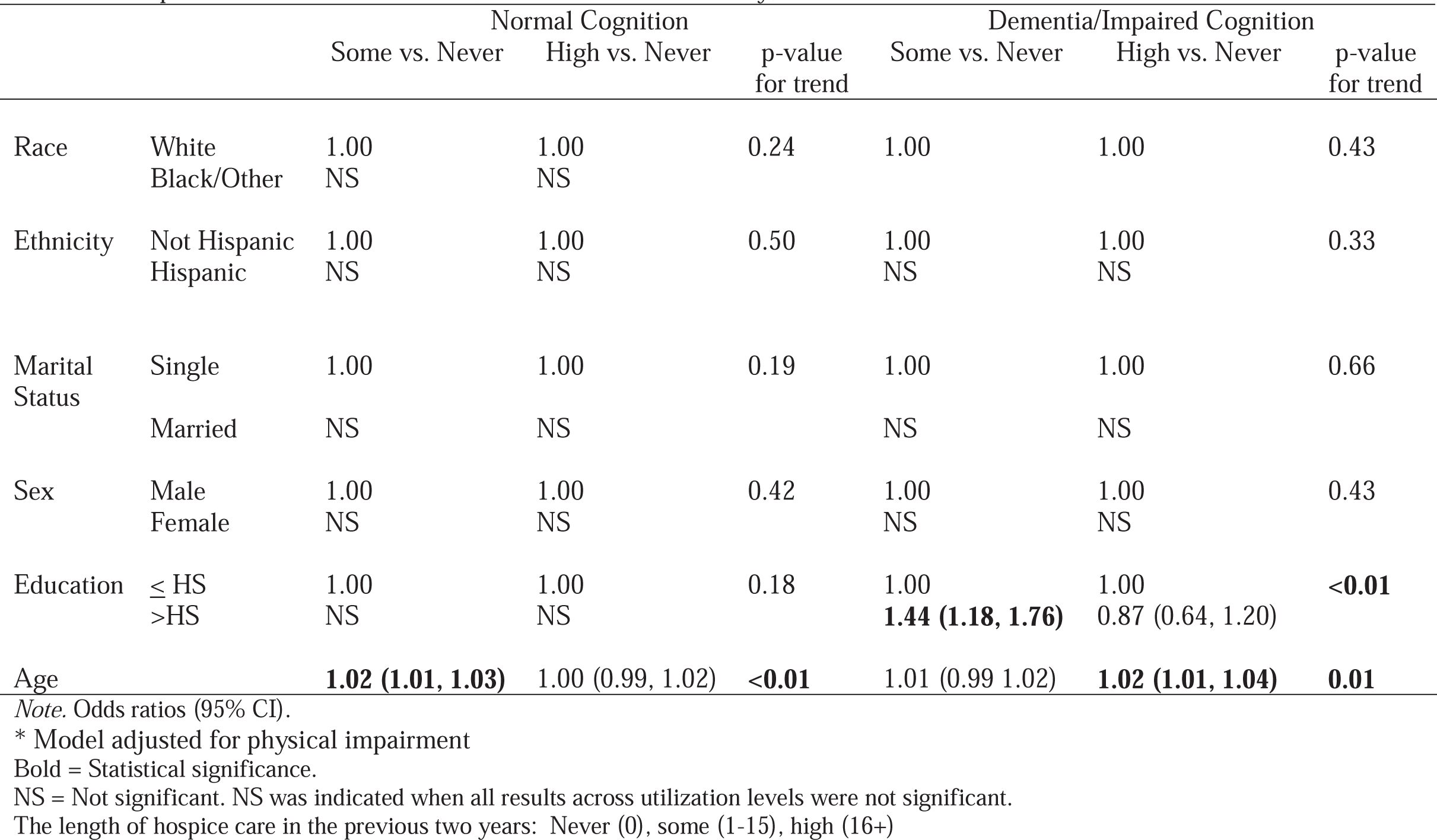
Hospice Care Prediction Model*, Health Retirement Survey, 2018.

### Doctor Visits

Among the normal cognition group, race, ethnicity, marital status, sex, education, and age had a significant trend across utilization levels of doctor visits (Table 6- trend test), with respondents from Black/Other race and Hispanic ethnicity less likely to experience doctors’ visits and those who are married, female, older and with greater levels of education were more likely to have doctors’ visits Among dementia/impaired cognition group, only ethnicity and education had significant trends in the utilization of doctor visits. higher education was associated with more doctor visits at all levels. Ethnicity also had a similar effect in both groups, with Hispanic participants less likely to visit the doctor at all frequencies assessed. (Table 6- trend test).

**Table 6.** Doctor’s Visits Prediction Model*, Health Retirement Survey, 2018.

|  |  | Normal Cognition |  |  |  | Dementia/Impaired Cognition |  |  |  |
| --- | --- | --- | --- | --- | --- | --- | --- | --- | --- |
|  |  | Low vs. Never | Moderate vs. Never | High vs. Never | p-value for trend | Low vs. Never | Moderate vs. Never | High vs. Never | p-value for trend |
| Race | White | 1.00 | 1.00 | 1.00 | <0.01 | 1.00 | 1.00 | 1.00 | 0.09 |
|  | Black/Other | **0.76 (0.67, 0.87)** | **0.67 (0.58, 0.78)** | **0.56 (0.44, 0.70)** |  | NS | NS | NS |  |
| Ethnicity | Not Hispanic | 1.00 | 1.00 | 1.00 | <0.01 | 1.00 | 1.00 | 1.00 | <0.01 |
|  | Hispanic | **0.35 (0.30, 0.40)** | **0.28 (0.23, 0.33)** | **0.20 (0.14, 0.27)** |  | **0.37 (0.29, 0.47)** | **0.32 (0.24, 0.42)** | **0.33 (0.20, 0.53)** |  |
| Rural | No | 1.00 | 1.00 | 1.00 | 0.08 | 1.00 | 1.00 | 1.00 | 0.85 |
|  | Yes | NS | NS | NS |  | NS | NS | NS |  |
| Marital Status | Single | 1.00 | 1.00 | 1.00 | <0.01 | 1.00 | 1.00 | 1.00 | 0.49 |
|  | Married | **1.30 (1.14, 1.49)** | **1.26 (1.09, 1.46)** | **1.39 (1.12, 1.72)** |  | NS | NS | NS |  |
| Sex | Male | 1.00 | 1.00 | 1.00 | <0.01 | 1.00 | 1.00 | 1.00 | 0.11 |
|  | Female | **1.63 (1.44, 1.85)** | **1.93 (1.68, 2.21)** | **2.00 (1.63, 2.45)** |  | NS | NS | NS |  |
| Education | ≤ HS | 1.00 | 1.00 | 1.00 | <0.01 | 1.00 | 1.00 | 1.00 | <0.01 |
|  | >HS | **1.75 (1.54, 1.99)** | **2.12 (1.85, 2.43)** | **1.82 (1.48, 2.22)** |  | **2.38 (1.80, 3.17)** | **2.90 (2.13, 3.93)** | **3.87 (2.50, 5.99)** |  |
| Age |  | **1.02 (1.01, 1.03)** | **1.04 (1.03, 1.05)** | **1.02 (1.01, 1.03)** | <0.01 | NS | NS | NS | 0.17 |

Note. Odds ratios (95% CI).
\* Model adjusted for number of chronic conditions and physical impairment.
Bold = Statistical significance.
NS = Not significant. NS was indicated when all results across utilization levels were not significant.
The number of doctor visits in the previous two years: never (0), low (1-6 visits), moderate (8-25 visits), and high (26+ visits).

### Mediation Analyses

We found no evidence for mediation analysis by our ACP measures for the associations identified between the SDOH variables and healthcare utilization previously noted (data not shown).

## Discussion

We evaluated healthcare utilization disparities among a nationally representative and diverse cohort of older adults with dementia and normal cognition in the context of SDOH (age, gender, race, ethnicity, education, marital status, and rural location). We found that compared to the normal cognition group, participants in the dementia/impaired cognition group were generally older, less educated, more likely to belong to racial/ethnic minority communities, more often single, and more likely to live in rural areas. Older adults with dementia also utilized a higher proportion of hospital, hospice, and nursing home care, had fewer low and medium doctor visits, and exhibited increased frequencies of both high and no doctor visits compared to those with normal cognition.

Overall, our findings revealed significant trends in healthcare utilization. Older age was associated with increased utilization across various healthcare services and cognition levels.

Hispanic ethnicity predicted fewer hospital stays in both groups and fewer doctor visits in the normal cognition group. Gender influenced hospital stays in dementia/impaired cognition group, nursing home stays in the normal cognition group, and doctor visits in both groups. Marital status was a predictor of nursing home stays and doctor visits in both groups. Race influenced nursing home stays in the dementia/impaired cognition group and doctor visits in the normal cognition group. Education predicted doctor visits in the dementia/impaired cognition group. No advance care planning measures mediated the associations between SDOH variables and healthcare utilization. Given the generally low rate of ACP among older adults in the U.S., further focused studies are needed to understand these results. More specifically, studies are needed to explore the barriers to ACP adoption and the potential impacts of increasing ACP engagement on healthcare outcomes.

Our analysis revealed several key trends in healthcare utilization. The Hispanic population tends to use less healthcare, likely due to lower access to healthcare services and other discriminatory factors (Mcmaughan et al., 2020; Rahemi & Williams, 2020). Older age was associated with higher healthcare use, which is understandable given the increase in age-related comorbidities. Married individuals used fewer long-term care services, such as nursing home stays, but had more outpatient care, such as doctor visits. The reduced use of long-term care services among married individuals may be attributed to the familial support they receive, which influences their preferences and allows them to remain at home for a longer period (Rahemi & Williams, 2016). Additionally, higher education significantly increased hospice care utilization, independent of advance care planning measures. This can underscore the influential role of education in healthcare decisions. We previously assessed the impact of SDOH on healthcare utilization in a 2014 cohort of HRS respondents; however, we did not account for ACP. We found that individuals in the impaired cognition/dementia group used more hospital and nursing home care, with either a high number of doctor visits or none at all and were less likely to have low to medium levels of visits compared to the normal cognition group (BLINDED FOR REVIEW). As demonstrated in the current study with the 2018 dataset, where we accounted for ACP, we observed that compared to the normal cognition group, participants in the dementia/impaired cognition group used more hospital, hospice, and nursing home care, had fewer low to medium doctor visits, and showed higher frequencies of both high and no doctor visits.

Our results align with prior research, suggesting that disparities in healthcare persist, particularly concerning socioeconomic factors like age, race/ethnicity, and education (Brown et al., 2016; Mcmaughan et al., 2020; Rahemi, Bacsu, et al., 2023; Rahemi, Malatyali, et al., 2023). In the scope of dementia, in particular, racial/ethnic minority populations are less likely to receive timely and accurate diagnoses, be prescribed anti-dementia medications, or use hospice care. They are also more likely to face a higher risk of hospitalization and receive more aggressive life-sustaining treatments, even at the end of life (Hinton et al., 2024). In a study using Medicare claims also, it was shown that Black beneficiaries faced higher risks of all-cause hospitalization, longer stays in skilled nursing facilities and shorter stays in hospice care compared to White beneficiaries. They were also less likely to receive physical/occupational therapy, dementia medications, and Parkinson’s disease medications (Lusk et al., 2023). These results highlight the disparity, especially considering that racial/ethnic minorities require more healthcare due to the disproportionate prevalence of dementia and comorbidities within these populations (Alzheimer’s Association, 2024; Charron-Chénier & Mueller, 2018).

As societies become increasingly multicultural and witness an unprecedented aging of their populations, the implications of our findings are significant. With the rising numbers of individuals affected by ADRD and the aging baby boomer population, continuous research on healthcare utilization in this group is critical. This research is vital for identifying trends, disparities, and developing tailored policies to meet the unique needs of diverse and minority communities. Our results underscore the importance of developing culturally tailored care management programs and policies that improve resource allocation in dementia care and advance care planning. By focusing on these areas and understanding the values and dignity of diverse older adult populations, we can work towards reducing health disparities and improving the overall quality of care for older adults with dementia.

## Strengths and Limitations

A key strength of this study is its use of a large, nationally representative sample of older adults from across the U.S. However, several limitations must be considered when interpreting the findings. First, secondary data analysis is constrained by the available data and measurement tools in the dataset, limiting researchers’ control. Second, the HRS study design results in an overrepresentation of African Americans, Hispanics, and Florida residents. To ensure accurate and unbiased estimates, we used survey weights for descriptive statistics but did not apply them in the modeling analyses, following established academic practices.

## Conclusion

Our study underscores the need for more research and policy development to address the healthcare disparities faced by an increasing number of diverse older adults, particularly those with dementia. As the aging population grows, it is crucial to understand the healthcare utilization patterns and disparities among diverse and minority communities. Our findings emphasize the importance of creating culturally tailored care management programs and policies that cater to the specific needs of these groups. By identifying factors that influence healthcare utilization, we can develop targeted interventions to reduce disparities and improve the quality of care for older adults with dementia. Respecting these populations’ values, desires, and dignity is vital for designing effective healthcare strategies and ensuring equitable access to healthcare services.

## Institutional Review Board (IRB) name and approval number

Clemson University Institutional Review Board exempted the study protocols (IRB Number: IRB2021- 0720).

## Data Availability

All data produced in the present work are contained in the manuscript

